# Risk of diabetes and cardiovascular diseases in women with vaginal bleeding before 20 gestational weeks: Danish population-based cohort study

**DOI:** 10.1101/2022.03.18.22272466

**Authors:** Elena Dudukina, Erzsébet Horváth-Puhó, Henrik Toft Sørensen, Vera Ehrenstein

## Abstract

**Importance:** The risk of cardiovascular outcomes following a pregnancy complicated by vaginal bleeding (VB) not ending in a miscarriage is not fully understood.

**Objective:** To study an association between pregnancy affected by VB and women’s subsequent risks of diabetes and cardiovascular outcomes.

**Design, Setting and Participants:** We conducted a population-based cohort study set in Denmark, 1994-2018. Using Danish nationwide registries, we identified 1,901,725 pregnancies among 903,327 women. Of these, 39,265 were affected by VB and ended in childbirth; 1,389,285 were unaffected by VB and ended in childbirth; 333,785 ended in termination, and 139,390 ended in a miscarriage.

**Exposure(s):** Pregnancy affected by vaginal bleeding before 20 gestational weeks.

**Main outcome measures:** The outcomes were diabetes types 1 and 2, hypertension, ischaemic heart disease (including myocardial infarction), atrial fibrillation or flutter, heart failure, ischaemic and haemorrhagic stroke. We computed absolute risks and hazard ratios (aHRs) with robust 95% confidence intervals (CIs) adjusted for age, calendar year, reproductive history, comorbidities, and socioeconomic factors using Cox proportional hazards regression.

**Results:** At the end of the follow-up, among women with VB-affected vs VB-unaffected pregnancies, the cumulative risks of the outcome events were 1.7% vs 1.4% for diabetes type 1; 6.9% vs 5.6% for diabetes type 2; 11.1% vs 9.0% for hypertension; 3.6% vs 2.7% for ischaemic heart disease, including 1.1% vs 0.8% for myocardial infarction; 1.3% vs 1.0% for atrial fibrillation or flutter; 0.5% vs 0.5% for heart failure; 2.0% vs 1.3% for ischaemic stroke; and 0.8% vs 0.6% for haemorrhagic stroke and aHRs were 1.2 to 1.3-fold increased for diabetes types 1 and 2, hypertension, ischaemic heart disease, myocardial infarction, atrial fibrillation or flutter, and heart failure. The aHRs were 1.4 and 1.5-fold increased for ischaemic and haemorrhagic stroke, respectively. For comparisons of women with VB-affected pregnancy vs termination, aHRs were up to 1.3-fold increased for diabetes and hypertension. Analyses contrasting VB-affected pregnancy with miscarriage resulted in aHRs below or close to the null value.

**Conclusions:** Women’s risks of diabetes and cardiovascular outcomes were increased following VB-affected pregnancy when compared with VB-unaffected pregnancy or termination, but not when compared with miscarriage.

**Key points:** *Question:* Is having a pregnancy affected by vaginal bleeding and not ending in miscarriage associated with the increased risk of diabetes and cardiovascular outcomes in women?

*Findings:* This nationwide registry-based cohort study of 1,901,725 pregnancies among 903,327 women showed that in contrast with having a pregnancy without vaginal bleeding, having a pregnancy with vaginal bleeding before 20 gestational weeks was associated with woman’s increased risks of diabetes types 1 and 2, hypertension, ischaemic heart disease, including myocardial infarction, atrial fibrillation or flutter, heart failure, ischaemic and haemorrhagic stroke. The cardiovascular risks were increased when investigating a woman’s first pregnancy with vaginal bleeding before 20 gestational vs without vaginal bleeding during pregnancy; the associations remained when we additionally adjusted for smoking and body-mass index in a sensitivity analysis.

*Meaning:* The results of this study support current clinical practice on cardiovascular prevention in women with pregnancy complications and add to the body of evidence on a common pregnancy complication, vaginal bleeding, and its association with long-term risks of diabetes and cardiovascular conditions.

## INTRODUCTION

Women’s reproductive health is a predictor of cardiovascular morbidity.^1^ First pregnancy ending in a miscarriage is associated with a 20% greater relative risk of hypertension and type 2 diabetes,^2,3^ whereas more than one miscarriage is associated with a 10-20% increased risk of stroke, thromboembolism, ischaemic heart disease, and 30% increased risk of cardiovascular diseases.^1,4,5^

Vaginal bleeding (VB) affects up to 30%^6–8^ of clinically recognised pregnancies a third of pregnancies complicated by VB ends in a pregnancy loss.^9,10^ Risk factors for VB in pregnancy are age ≥ 35 years, obesity, lack of physical exercise, stress, cigarette smoking, alcohol abuse, inflammation, and low plasma levels of progesterone during gestation.^9,11–18^ There are little data on whether VB not followed by a pregnancy loss is associated with subsequent cardiovascular morbidity similarly to miscarriage. An earlier Danish study^19^ reported a 25-60% higher risk of multiple cardiovascular outcomes in women following a pregnancy with VB in the first trimester without subsequent miscarriage vs unaffected pregnancy; however, analyses did not control for pre-pregnancy morbidity and socioeconomic factors. Comparisons of VB-affected pregnancies with pregnancies ending in terminations or miscarriages are missing in the literature.

We investigated the association of VB before 20 gestational weeks of a pregnancy ending in a childbirth with the woman’s subsequent risks of diabetes, hypertension, ischaemic heart disease, including myocardial infarction, atrial fibrillation or flutter, heart failure, and ischaemic and haemorrhagic stroke when compared with having a VB-unaffected pregnancy, termination, or miscarriage.

## METHODS

### Setting and design

We conducted a registry-based cohort study using prospectively and routinely recorded data of the Danish nationwide administrative and health registries,^20,21^ which are collected in the context of Denmark’s universal tax-supported healthcare system. Data from all registries are linkable via a unique personal identifier stored in the Civil Registration System (CRS).^20^ It captures the civil and vital status of all residents, country of origin, immigration, and emigration dates. Data on all deliveries are stored in the Medical Birth Registry (MBR) available from 1973.^22^ Smoking and body mass index (BMI) in pregnancy became available in the MBR starting in 1991 and 2004, respectively. Hospital encounters and procedures are captured by the Danish National Patient Registry (DNPR).^23^ It stores data on medical procedures, inpatient hospital encounters since 1977 and also on outpatient specialist clinic visits and emergency room visits starting in 1995.^23^ The diagnoses recorded in the DNPR are coded according to the International Classification of Diseases, 8^th^ revision (ICD-18) in 1977-1993 and 10^th^ revision (ICD-10) from 1994 onwards, while procedures are coded using the Danish Classification of Surgical Procedures and Therapies before 1996 and Nordic Medico-Statistical Committee Classification of Surgical Procedures afterwards.^23^ The psychiatric admissions and outpatient specialist clinic encounters were available for this study starting from 1995 as a part of the DNPR.^24^ Nationwide data on medication prescriptions were available starting in 1995 from the Danish National Prescription Registry.^25^ This registry captures the Anatomical Therapeutic Classification code of the prescribed medication, and the date a prescription was redeemed.^25^ Data on education,^26^ employment^27^ and income^28^ are available from 1980, measured yearly and stored at Statistics Denmark.^29^

### Study population

From the MBR, we assembled a cohort of women whose pregnancies ended in a live or stillbirth. Each delivery of multi-foetal pregnancy was counted as one pregnancy (Figure 1). From the DNPR, we ascertained two cohorts of women whose pregnancies ended in a termination or miscarriage.

**Figure 1.**
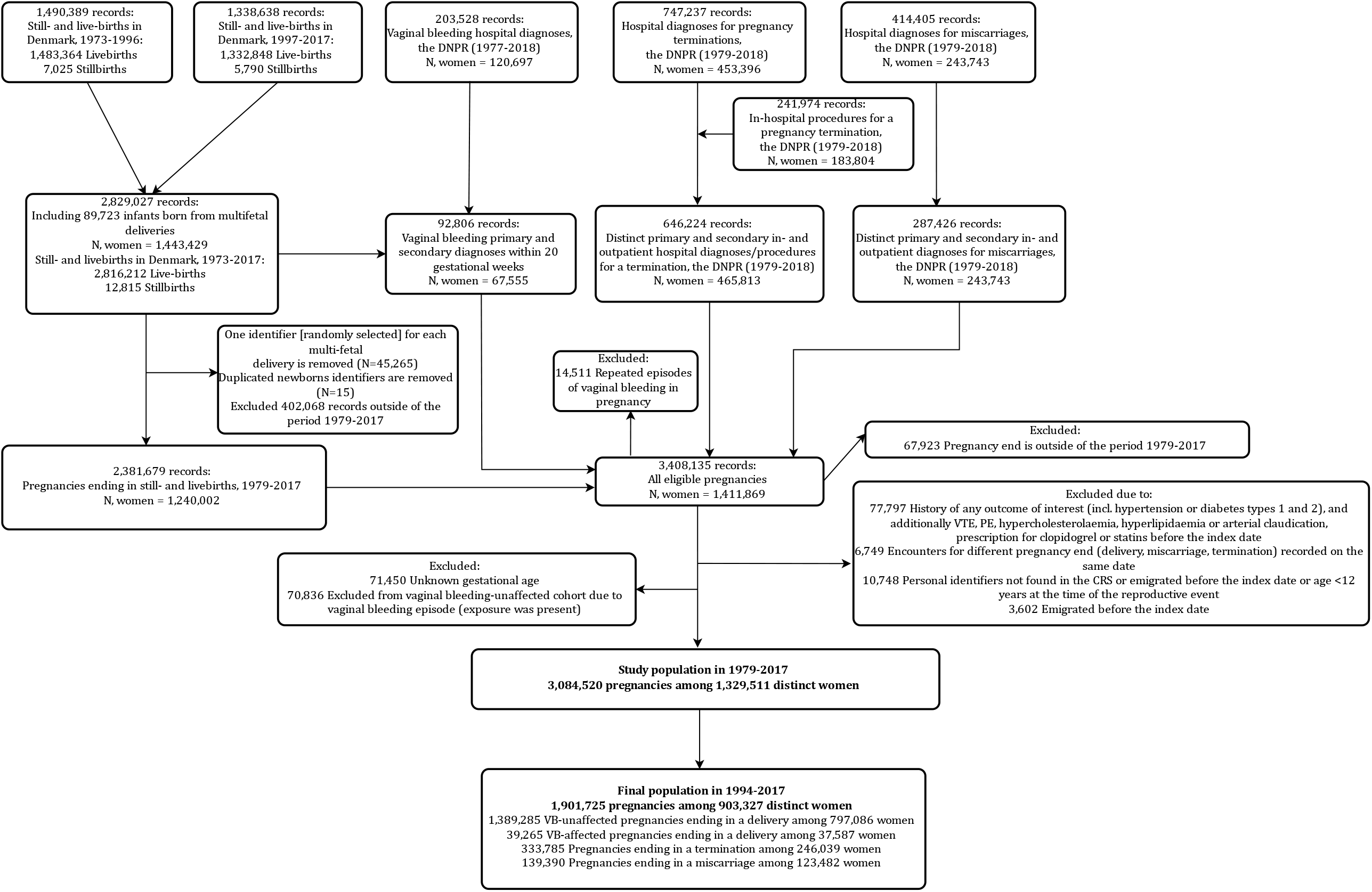
Flow graph depicting ascertainment of the study population. Standalone file

We excluded records of women with any pre-existing outcome of interest, venous thrombotic event, records of women with two or more prescriptions for antihypertensives from different classes redeemed within 180 days of each other before the index date as a proxy for hypertension as well as records of women with pre-existing hypercholesterolaemia, hyperlipidaemia or arterial claudication, prescription for clopidogrel or statins before the index date (altogether 2.3% of all eligible observations). We further excluded records with missing gestational age (2.1% of all eligible observations) and records with a delivery, miscarriage, termination, or VB recorded on the same date (0.2% of all eligible observations). Finally, we excluded records of women whose personal identifier could not be linked with the CRS data, records with potentially wrongly registered women’s age (<12 years) at the time of hospital-based encounter for delivery, miscarriage, termination, or VB (0.2% of all eligible observations); and pregnancies recorded after a woman emigrated from Denmark (0.1% of all eligible observations) (Figure 1). We also ascertained the subset of women having their first pregnancy recorded as either delivery, miscarriage or termination.

### Exposed cohort

Among women having a pregnancy ending in a childbirth, we identified pregnancies affected by VB before 20 gestational weeks, using the DNPR. When several VB episodes occurred before 20 gestational weeks of the same pregnancy, the first episode served as the qualifying event (Figure 1). The index date was the date of delivery when all cohort-specific eligibility criteria were met.

### The comparison cohorts

The women whose pregnancies ended in a delivery and had no record of VB before the 20th gestational week in the DNPR formed a comparison cohort of women with VB-unaffected pregnancy and the index date was the date of delivery.

We ascertained women having a pregnancy ending in a termination or miscarriage using the primary and secondary diagnoses at any hospital visit. When a miscarriage or pregnancy termination encounter had several dates associated with it (hospital admission, procedure, hospital discharge), we selected the earliest of them as the index date (Table 1 in the Supplementary materials). A woman with a pregnancy termination encounter characterised by the same diagnostic or procedure code as her previous encounter could re-enter the population with another pregnancy after 180 days. We assumed that the hospital encounter with the same pregnancy-related diagnostic or procedure code within 180 days since an earlier such encounter may be related to the complications of the previous event. We applied the same algorithm for women with pregnancies ending in miscarriage.

**Table 1.**
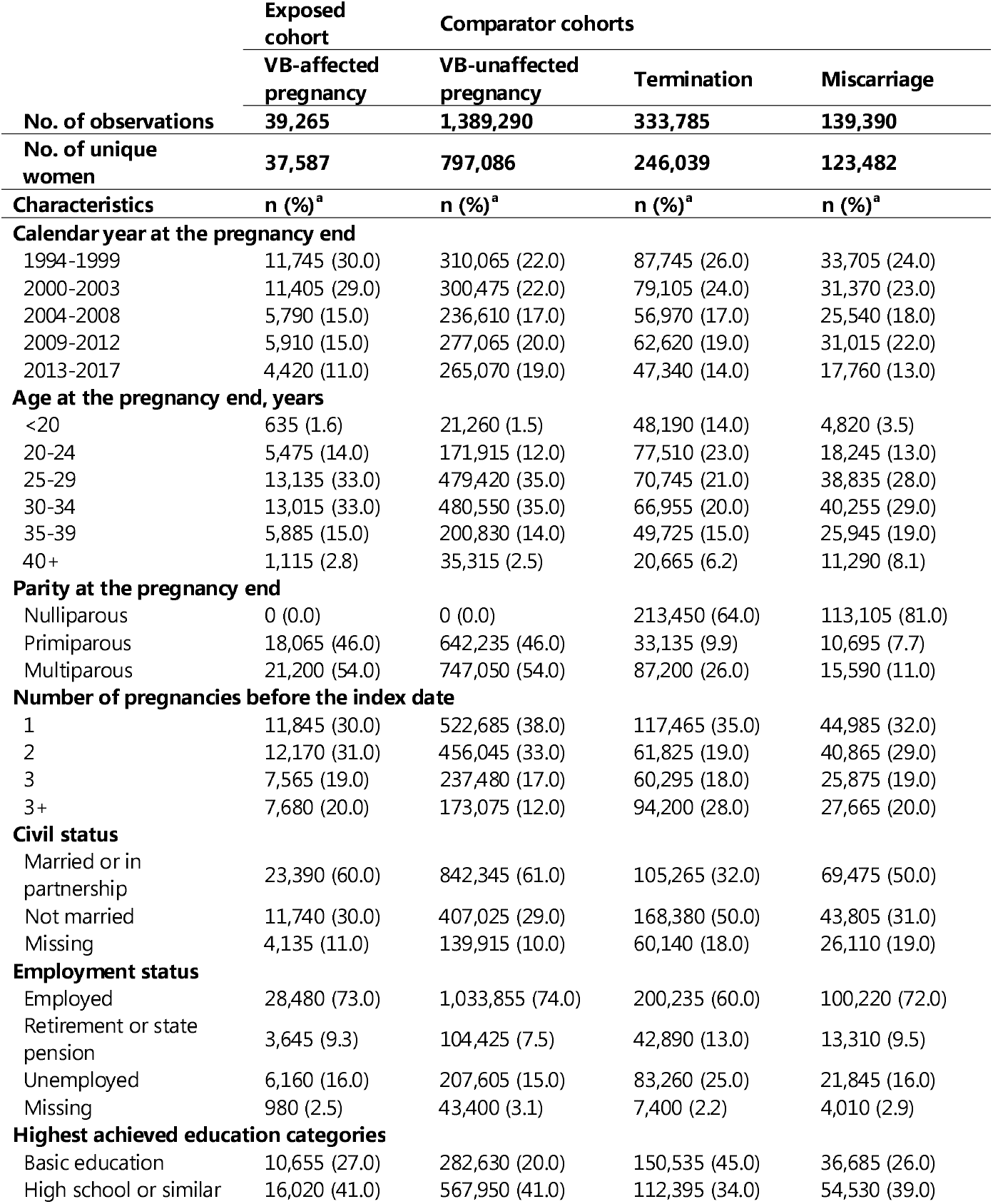
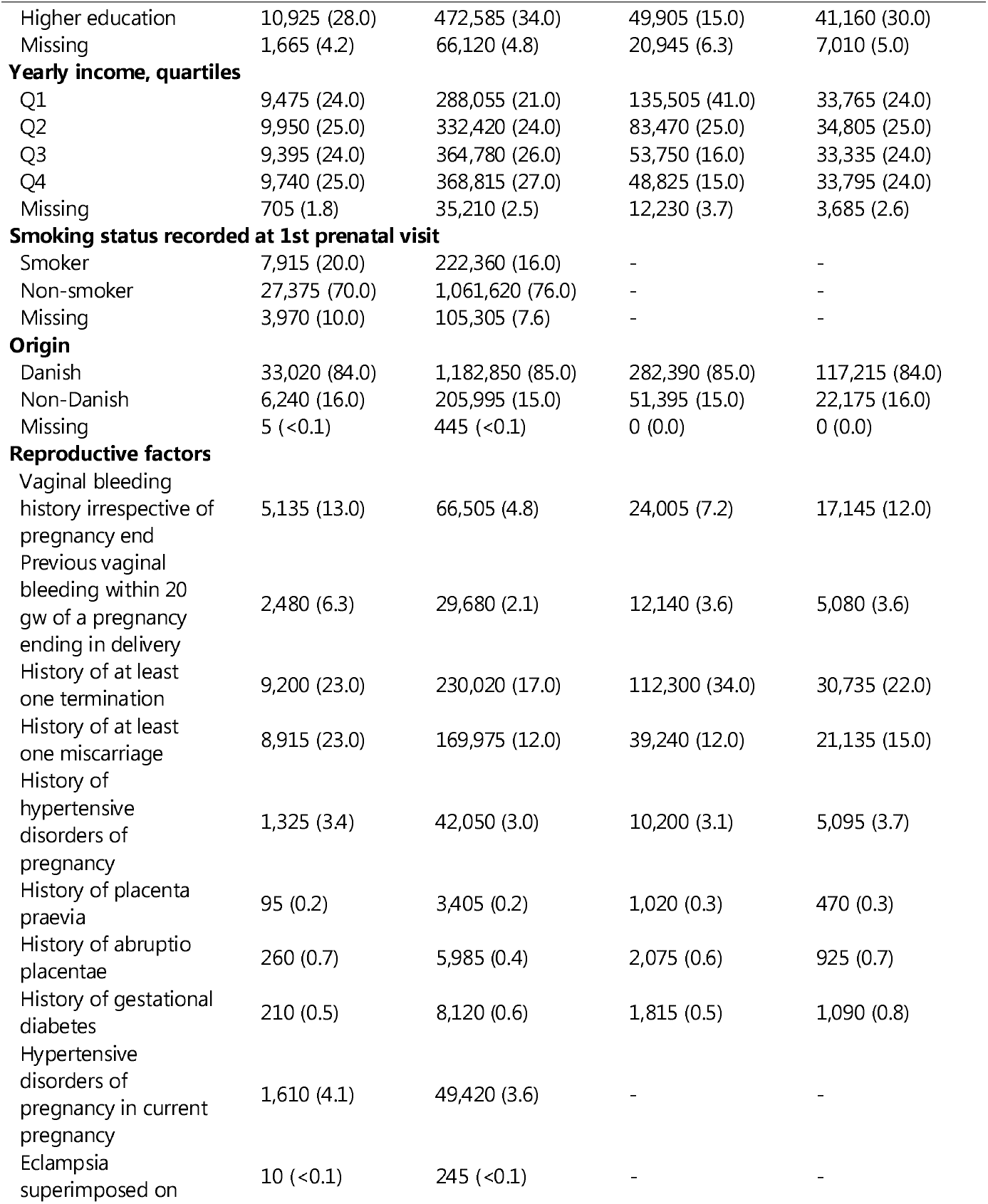
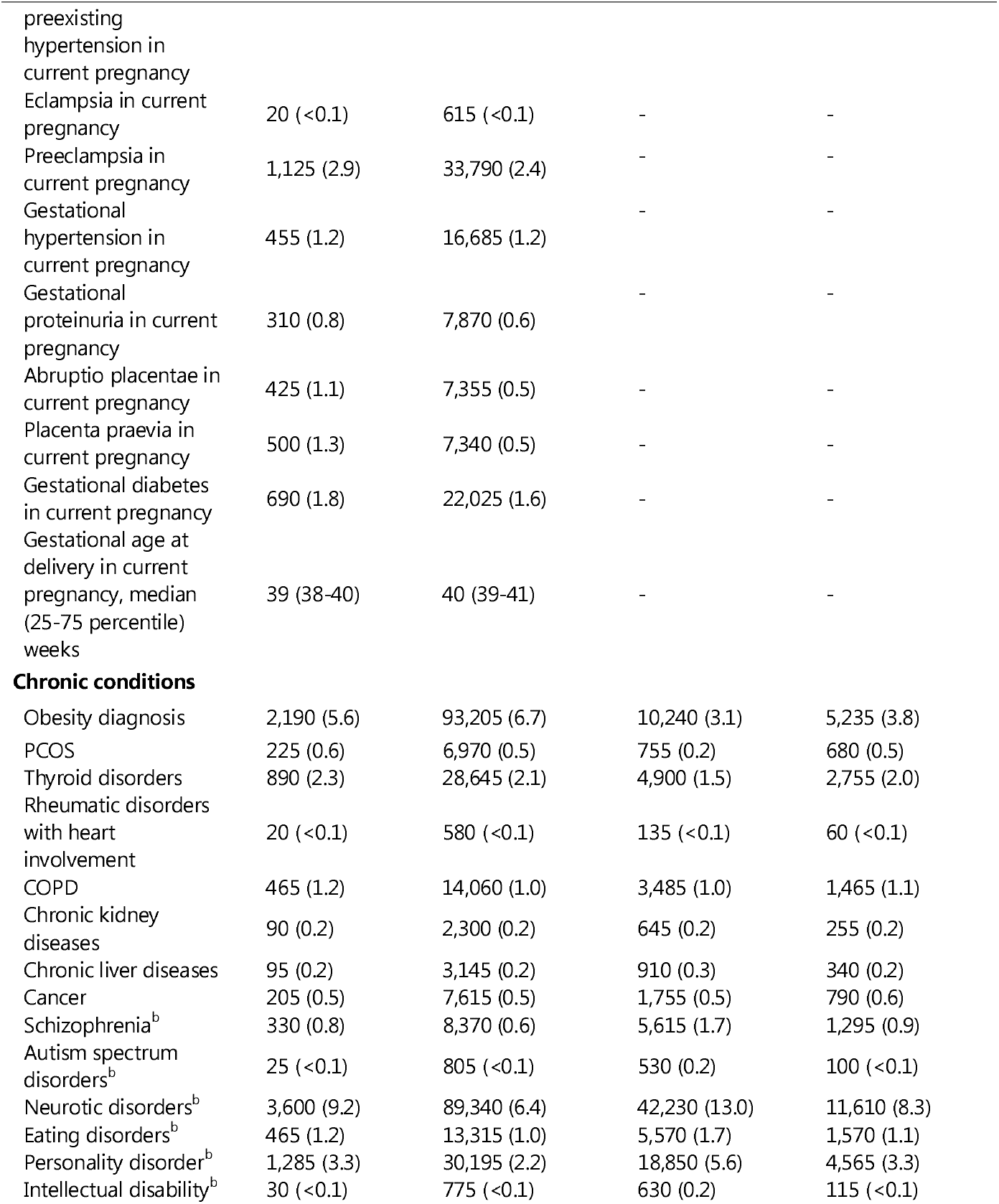
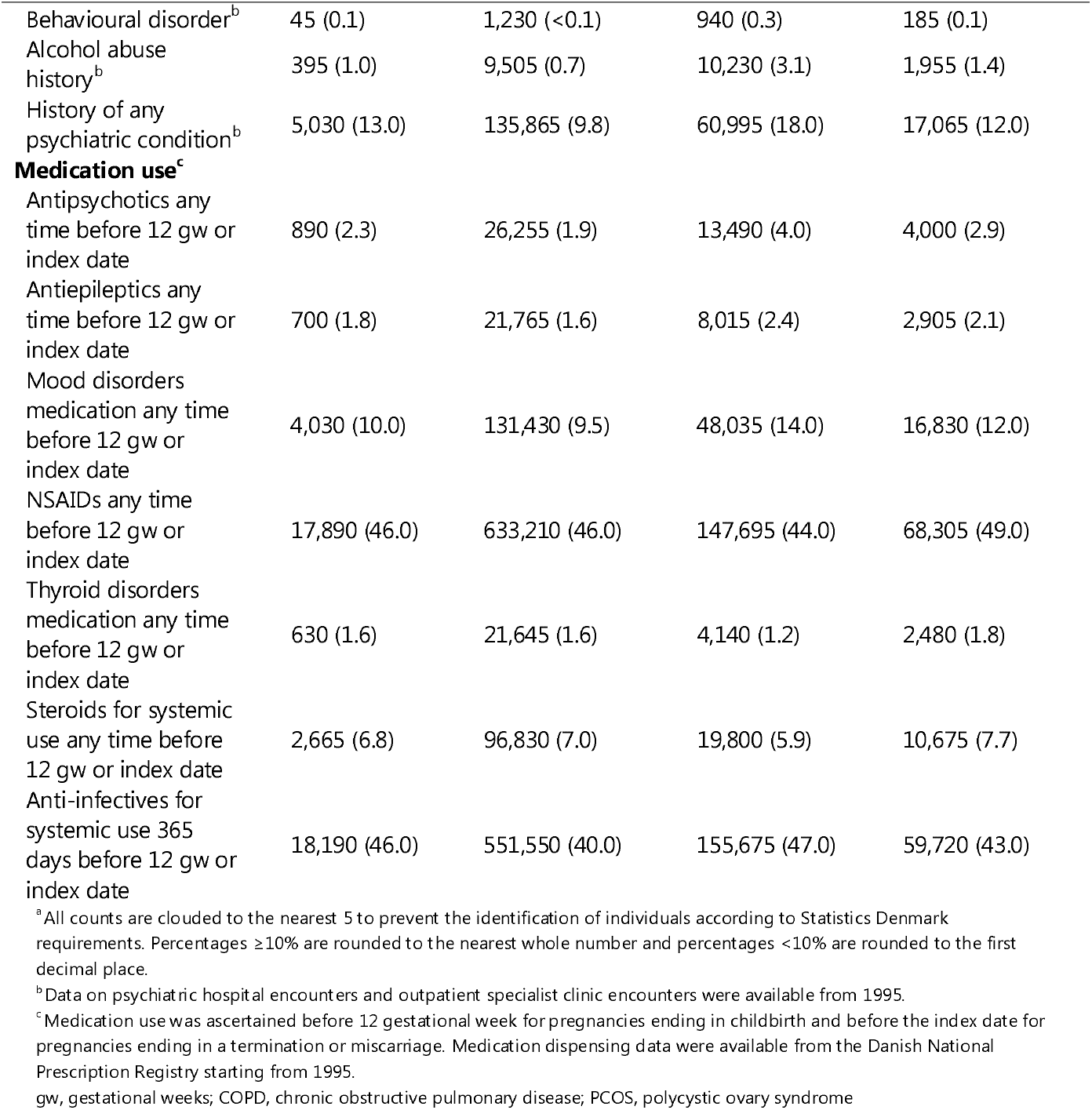
Descriptive characteristics of vaginal bleeding-affected pregnancy cohort and comparator cohorts, Denmark, 1994-2017.

### Outcomes

The outcomes were incident diabetes type 1 and type 2, hypertension, ischaemic heart disease, including myocardial infarction, atrial fibrillation or flutter, heart failure, ischaemic and haemorrhagic stroke. We used all available primary and secondary diagnoses recorded in connection with inpatient, outpatient specialist clinic, and emergency room encounters from the DNPR (1994-2018). Diabetes and hypertension outcomes were defined based on the earliest of the hospital-based diagnosis or medication dispensing (Table 1 in the Supplementary materials). In a sensitivity analysis with the follow-up from 1979 through 2018, we included two additional long-term cardiovascular outcomes (coronary artery bypass grafting [CABG] and percutaneous coronary intervention [PCI]). These outcomes were defined using procedure codes from the DNPR (Table 1 in the Supplementary materials).

### Covariables

Potential confounding factors were selected using the directed acyclic graph presenting the evidence on the associations or lack thereof between the measured and unmeasured variables in this study (Figure 1 in the Supplementary materials). These were women’s age, calendar year of pregnancy end, parity (nulli- or primiparous vs multiparous) as the number of delivered live and stillborn infants, pre-index date number of identifiable pregnancies, civil status, employment status, highest attained education, annual women’s income in quartiles; reproductive history (VB diagnosis irrespective of a later delivery, history of VB before 20th gestational week of pregnancy ending in a childbirth, histories of at least one termination or miscarriage, history of preeclampsia-eclampsia, placenta praevia, abruptio placentae, gestational diabetes), obesity, chronic obstructive pulmonary disease (COPD), chronic kidney and liver diseases, cancer, rheumatic conditions with heart involvement, and psychiatric conditions before the index date. As a proxy for the underlying long-term health, we also ascertained the medication use (antipsychotics, mood disorders medication, antiepileptics, non-steroid anti-inflammatory drugs [NSAIDs], steroids for systemic use, anti-infectives) history before the end of 12^th^ gestational week for pregnancies ending in a childbirth and before the index date for pregnancies ending in termination or miscarriage.

### Statistical analysis

We followed each identified pregnancy of a woman from its index date until the earliest of each of the outcomes, emigration, death, or end of data on 31 December 2018. We constructed the cumulative incidence curves for the outcomes while treating death and emigration as competing risks^30^ and computed incidence rates (IR) of the outcomes per 10,000 person-years (PY) for women having VB-affected and VB-unaffected pregnancy, termination, or miscarriage. We used Cox proportional hazards regression to compute the adjusted hazards ratios (HRs) for the outcomes of interest with the corresponding 95% confidence intervals (CIs) adjusted for age, calendar year, reproductive history, comorbidities, and socioeconomic factors. In analyses of all identifiable pregnancies, for each subsequent pregnancy, we took into account exposure status at previous pregnancy as a covariable. Different pregnancies of the same woman could contribute person-time to multiple cohorts. Different outcomes did not censor one another. In analyses of all identifiable pregnancies, we did not censor an observation at the time the next pregnancy occurred to avoid informative censoring. To account for different pregnancies in the same woman, we computed the 95% CIs accounting for multiple non-independent observations of the same woman as incorporated in *survival* and *WeightIt* R packages.^31–34^

We computed the inverse probability of treatment (IPT) weights for VB-affected pregnancies vs each of the comparator cohorts.^34–36^ We trimmed large IPT weights (>50) at the 99^th^ percentile.^34,36^ The covariables balance after IPT weighting was evaluated using standardized mean difference (SMD). Finally, we repeated all analyses restricting to a woman’s first identifiable pregnancy. Additionally, we computed conventionally adjusted HRs using multivariable Cox proportional hazards regression models. We evaluated the proportionality of hazards assumption by using log-minus-log plots and found no violations.

We performed several sensitivity analyses. First, we conducted analyses of women with identifiable pregnancies in the ICD-8 era from 1979 through 1993 and investigated the risks of two additional outcomes (CABG and PCI) with the follow-up through 2018. Second, we restricted analyses to women with pregnancies from 2004 through 2017 while further adjusting analyses of VB-affected vs VB-unaffected pregnancies for pre-pregnancy BMI. Third, we investigated the association between VB and myocardial infarction and stroke outcomes restricted to inpatients with an acute admission. For the contrasts of VB-affected vs VB-unaffected pregnancy, we computed E-values quantifying the strength of the unmeasured confounding needed to explain away the findings.

For all analyses, we used RStudio^37^ and R^38^ versions 3.6.0-4.1.0 including the following packages and their dependencies: tidyverse,^39^ ggplot2,^40(p2)^ survival,^32^ cmprsk,^41^ epiR,^42^ and WeightIt (Weighting for Covariate Balance in Observational Studies).^34^ All presented counts were masked to comply with the rules of Statistics Denmark.^29^ We did not impute missing values for covariables since the missingness was relatively rare.

No patient informed consent or ethical approval is required for registry-based research according to Danish legislation. The study was reported to the Danish Data Protection Agency^43^ through registration at Aarhus University (record number: AU-2016-051-000001, sequential number 605).

## RESULTS

### Descriptive characteristics

Between 1994 and 2017, we identified 1,901,725 pregnancy records among 903,327 women. Of ascertained pregnancies, 1,428,550 ended in a delivery (39,265 [2.1%] were VB-affected before 20^th^ week of gestation and 1,389,285 [73.1%] were VB-unaffected pregnancies); 333,785 (17.6%) ended in a termination, and 139,390 (7.2%) pregnancies ended in a miscarriage.

Women having VB-affected and VB-unaffected pregnancies had a similar distribution of age at delivery, parity, civil status, employment, highest attained education, yearly income, history of COPD, obesity, polycystic ovary syndrome (PCOS), chronic kidney and liver disease, alcohol abuse, and medication use (Table 1). Women having VB-affected pregnancies were more likely than women having VB-unaffected pregnancies to have a prior miscarriage (23.0% vs 12.0%), prior VB-affected pregnancy (6.3% vs 2.1%) and a prior psychiatric condition (13.0% vs 9.8%). For deliveries at investigation, VB-affected vs VB-unaffected pregnancies were more often complicated by abruption placentae (1.1% vs 0.5%) and placenta praevia (1.3% vs 0.5%). Women with a termination were more likely to be younger, nulliparous, not married, in the first income quartile and to have a prior psychiatric condition (Table 1). The median follow-up (25-75^th^ percentile) was 13.3 (7.2-19.2) years and differed slightly by the outcome at the investigation. The main analyses accrued 593,034 PY for VB-affected cohort, 18,170,486 PY for VB-unaffected cohort, 4,662,913 PY for terminations, and 1,917,129 PY for miscarriages.

### Risks and rates of the outcomes

Following VB-affected pregnancy there were 365 events of diabetes type 1 (IR=6.1, 95% CI: 5.5-6.8 per 10,000 PY); 1,425 events of diabetes type 2 (24.4, 95% CI: 23.1-25.6); 2,120 events of hypertension (36.5, 95% CI: 34.9-38.0); 720 events of ischaemic heart disease (12.2, 95% CI: 11.4-13.1), including 180 events of myocardial infarction (3.0, 95% CI: 2.6-3.5); 235 events of atrial fibrillation or flutter (4.0, 95% CI: 3.5-4.5); 100 events of heart failure (1.7, 95% CI: 1.4-2.0); 390 ischaemic stroke events (6.6, 95% CI: 5.9-7.2) and 165 haemorrhagic stroke events (2.8, 95% CI: 2.4-3.3) (Table 2).

**Table 2.**
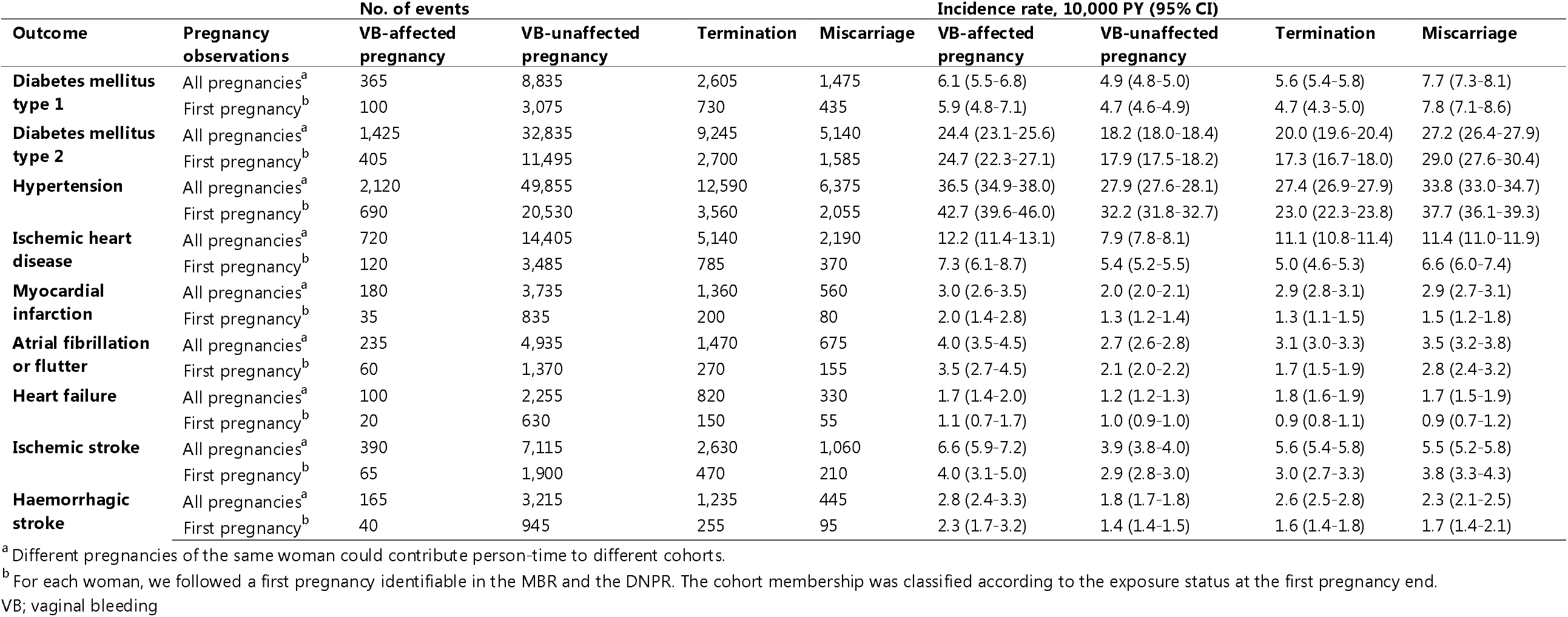
Incidence rates of diabetes and cardiovascular diseases following vaginal bleeding-affected pregnancy cohort and comparator cohorts, Denmark, 1994-2017.

At the end of follow-up, for women with VB-affected vs VB-unaffected pregnancies, the cumulative incidences of the outcomes were: 1.7% vs 1.4% for diabetes type 1; 6.9% vs 5.6% for diabetes type 2; 11.1% vs 9.0% for hypertension; 3.6% vs 2.7% for ischaemic heart disease, including 1.1% vs 0.8% for myocardial infarction; 1.3% vs 1.0% for atrial fibrillation or flutter; 0.5% vs 0.5% for heart failure; 2.0% vs 1.3% for ischaemic stroke; and 0.8% vs 0.6% for haemorrhagic stroke (Figure 2 in the Supplementary materials). The cumulative incidences of the outcomes at 5, 10, 20, and 25 years of follow-up are reported in Table 2 in the Supplementary materials.

### Hazard ratios

After IPT weighting and trimming, all covariables were balanced within SMD cut-off of 0.05 for women with VB-affected vs VB-unaffected pregnancy and vs miscarriage and within SMD cut-off of 0.15 for women with VB-affected pregnancy vs pregnancy ending in a termination.

Women with VB-affected vs VB-unaffected pregnancy, were at a greater risk of diabetes type 1 (IPTW-adjusted HR [IPTW aHR]: 1.15, 95% CI: 1.03-1.28), diabetes type 2 (IPTW aHR: 1.19, 1.13-1.26), hypertension (IPTW aHR: 1.19, 1.14-1.25), ischaemic heart disease (IPTW aHR: 1.26, 1.16-1.37) and specifically myocardial infarction (IPTW aHR: 1.21, 1.03-1.42), atrial fibrillation or flutter (IPTW aHR: 1.32, 1.14-1.51), heart failure (IPTW aHR: 1.23, 0.99-1.52), ischaemic (IPTW aHR: 1.41, 1.26-1.57) and haemorrhagic stroke (IPTW aHR: 1.46, 1.23-1.72). The elevated risks persisted in the analyses following a woman’s first pregnancy (Table 2). The conventionally adjusted HRs were similar in magnitude for all outcomes but heart failure (conventionally aHR: 1.08, 0.88-1.32; Table 3 in the Supplementary materials). When comparing women with VB-affected pregnancy vs having a termination, the risks were increased for diabetes type 1 (IPTW aHR: 1.29, 95% CI: 1.11-1.49), diabetes type 2 (IPTW aHR: 1.33, 1.23-1.43), hypertension (IPTW aHR: 1.35, 1.26-1.43), ischaemic heart disease (IPTW aHR: 1.20, 1.07-1.33), atrial fibrillation or flutter (IPTW aHR: 1.12, 0.93-1.36), ischaemic stroke (IPTW aHR: 1.14, 0.98-1.32). Associations with myocardial infarction (IPTW aHR: 1.12, 0.90-1.39) and heart failure (IPTW aHR: 1.12, 0.85-1.49) were compatible with the null value and showed estimates close to unity in conventionally adjusted analyses. In analyses of a woman’s first pregnancy, the associations were 1.2 to 1.4-fold increased for diabetes types 1 and 2, hypertension, ischaemic heart disease, and ischaemic and haemorrhagic stroke (Table 3).

**Table 3.** Adjusted hazard ratios with 95% confidence intervals for the associations of vaginal bleeding-affected pregnancy with diabetes and cardiovascular diseases, Denmark, 1994-2018.

| Outcomes | VB-affected pregnancy vs<br>Comparator | Pregnancy observations |  |
| --- | --- | --- | --- |
|  |  | All<br>observations<sup>a</sup> | First<br>observations<sup>b</sup> |
|  |  | HR (95% CI)<sup>c</sup> |  |
| Diabetes mellitus type 1 | VB-unaffected pregnancy<sup>d</sup> | 1.15 (1.03-1.28) | 1.11 (0.98-1.26) |
|  | Termination | 1.29 (1.11-1.49) | 1.25 (1.07-1.45) |
|  | Miscarriage | 0.81 (0.69-0.95) | 0.85 (0.74-0.98) |
| Diabetes mellitus type 2 | VB-unaffected pregnancy<sup>d</sup> | 1.19 (1.13-1.26) | 1.30 (1.22-1.39) |
|  | Termination | 1.33 (1.23-1.43) | 1.37 (1.27-1.49) |
|  | Miscarriage | 0.86 (0.80-0.94) | 0.98 (0.91-1.06) |
| Hypertension | VB-unaffected pregnancy<sup>d</sup> | 1.19 (1.14-1.25) | 1.16 (1.11-1.21) |
|  | Termination | 1.35 (1.26-1.43) | 1.16 (1.10-1.22) |
|  | Miscarriage | 1.09 (1.02-1.17) | 0.99 (0.94-1.04) |
| Ischemic heart disease | VB-unaffected pregnancy<sup>d</sup> | 1.26 (1.16-1.37) | 1.37 (1.26-1.49) |
|  | Termination | 1.20 (1.07-1.33) | 1.19 (1.08-1.32) |
|  | Miscarriage | 0.96 (0.86-1.09) | 1.07 (0.97-1.17) |
| Myocardial infarction | VB-unaffected pregnancy<sup>d</sup> | 1.21 (1.03-1.42) | 1.24 (1.05-1.47) |
|  | Termination | 1.12 (0.90-1.39) | 1.03 (0.84-1.26) |
|  | Miscarriage | 0.85 (0.67-1.09) | 0.96 (0.80-1.16) |
| Atrial fibrillation or<br>flutter | VB-unaffected pregnancy<sup>d</sup> | 1.32 (1.14-1.51) | 1.26 (1.10-1.45) |
|  | Termination | 1.12 (0.93-1.36) | 0.86 (0.72-1.02) |
|  | Miscarriage | 1.01 (0.83-1.23) | 0.85 (0.73-0.99) |
| Heart failure | VB-unaffected pregnancy<sup>d</sup> | 1.23 (0.99-1.52) | 1.15 (0.94-1.42) |
|  | Termination | 1.12 (0.85-1.49) | 0.83 (0.65-1.06) |
|  | Miscarriage | 0.89 (0.65-1.20) | 0.83 (0.66-1.05) |
| Ischemic stroke | VB-unaffected pregnancy<sup>d</sup> | 1.41 (1.26-1.57) | 1.30 (1.15-1.47) |
|  | Termination | 1.14 (0.98-1.32) | 1.07 (0.92-1.25) |
|  | Miscarriage | 0.93 (0.79-1.10) | 1.02 (0.89-1.17) |
| Haemorrhagic stroke | VB-unaffected pregnancy<sup>d</sup> | 1.46 (1.23-1.72) | 1.34 (1.12-1.61) |
|  | Termination | 1.10 (0.87-1.38) | 1.18 (0.94-1.48) |
|  | Miscarriage | 0.96 (0.75-1.23) | 1.12 (0.91-1.38) |

<sup>a</sup> Different pregnancies of the same woman could contribute person-time to different cohorts.
<sup>b</sup> We followed a first pregnancy of each woman identifiable via the MBR and the DNPR. The cohort membership was classified according to the exposure status at the first pregnancy.
<sup>c</sup> Adjusted using IPT-weighted Cox proportional hazards regression for women's age at pregnancy end, calendar year, parity, number of previous pregnancies ending in childbirth, termination or miscarriage and identifiable via the MBR and the DNPR, civil status, employment, highest attained education, income in year-specific quartiles, reproductive history (previous vaginal bleeding, termination, miscarriage, preeclampsia-eclampsia, placenta praevia, abruption placentae), comorbidities (obesity, chronic pulmonary disease, chronic kidney disease, chronic liver disease, cancer, rheumatic conditions with heart involvement, thyroid conditions, alcohol abuse), history of any psychiatric condition, and medication use (antipsychotics, antiepileptics, medication for mood disorders, NSAIDs or aspirin, thyroid disorders medication, steroids for systemic use, and anti-infectives for systemic use).
<sup>d</sup> Analyses were additionally adjusted for smoking status recorded at 1st prenatal visit, preeclampsia-eclampsia, placenta praevia, and abruption placentae in the index pregnancy.
CI, confidence interval; HR, hazard ratio

When contrasted with having a miscarriage, having a VB-affected pregnancy was not associated with hypertension, ischaemic heart disease, including myocardial infarction, atrial fibrillation or flutter, heart failure, ischaemic, and haemorrhagic stroke in women. IPTW aHRs for diabetes types 1 and 2 suggested reduced risks of these outcomes (Table 3).

In the comparisons of VB-affected vs VB-unaffected pregnancy and when investigating pregnancies with the index date in 1979-1993, the risks remained increased for all outcomes after additional adjustment for women’s BMI. In these analyses, for the contrast of VB-affected pregnancies vs VB-unaffected pregnancies, the IPTW aHR were 1.28 (95% CI: 0.98-1.68) and 1.20 (95% CI: 1.06-1.35) for CABG and PCI, respectively (Table 4 and Table 5 in the Supplementary materials). Results of analyses for acute myocardial infarction, ischaemic and haemorrhagic stroke were similar to the results of the main analyses; however, the estimates for comparisons of VB-affected vs VB-unaffected pregnancies shifted slightly further away from the null (Table 6 in the Supplementary materials).

An unmeasured confounder associated with VB and with cardiovascular outcomes with the hazard ratio of 1.6-2.2 beyond measured covariables could fully explain away the observed associations according to the E-value analysis (Table 7 in the Supplementary materials).^44^

## DISCUSSION

### Principal findings

In this population-based cohort study, the hazards of diabetes types 1 and 2 as well as of multiple cardiovascular outcomes were 1.2 to 1.5-fold increased for women with VB-affected vs VB-unaffected pregnancies. The associations persisted in the analyses restricted to first identifiable pregnancies and in the sensitivity analyses additionally controlling for BMI and allowing for a longer follow-up. We found that the hazards of diabetes types 1 and 2, hypertension, ischaemic heart disease, atrial fibrillation or flutter, and haemorrhagic stroke were 1.2 to 1.3-fold greater for women having a VB-affected pregnancy vs termination. No increased risks of the outcomes of interest were observed in the comparisons of women having a VB-affected pregnancy vs miscarriage. The findings suggest that having a pregnancy complicated by VB was associated with increased cardiovascular risk.

### Biological mechanisms

Underlying biological mechanisms of VB in pregnancy are not fully known. Markers of systemic inflammation are associated with both miscarriages^45,46^ and VB in pregnancy^47–50^ and were found to contribute to the development of cardiovascular disease and diabetes type 2.^51^ Dyslipidaemia, insulin resistance, and obesity are risk factors for diabetes type 2 and hypertension, which result in endothelial dysfunction and microvascular inflammation and damage further leading to cardiovascular diseases.^52^ The existence of common metabolic and pro-inflammatory pathways via cytokines (IL-1β, IL-6, TNF-α) for VB, miscarriage and vascular pathology could contribute to the explanation of the findings of this study showing an elevated cardiovascular risk in women with a VB-affected pregnancy vs women having a VB-unaffected pregnancy or a termination but not a miscarriage. Of note, type 1 diabetes mellitus has a distinct aetiology related to autoimmune or non-autoimmune destruction of insulin-producing β-cells.^53^ This may support that VB aetiology has pronounced pro-inflammatory pathways; however, it may also point to the residual confounding in this study.

### Comparison with other studies

An earlier Danish study showed a greater risk of cardiovascular morbidity following pregnancies with VB.^19^ This study differed from the earlier study in several ways. First, we used a different definition of VB and aligned it with the clinical definition of threatened miscarriage, more years of follow-up were available for this study and investigated outcomes included specific stroke and diabetes types. Second, we carried out comparisons with pregnancies ending in a termination or miscarriage. Third, analyses in this study were adjusted for socioeconomic factors and pre-pregnancy morbidity. We did not adjust for post-exposure factors such as preterm delivery, prelabour rupture of membranes, foetal growth restriction, placental abruption and stillbirth. These could be both mediators and colliders on the pathway from VB in pregnancy to subsequent cardiovascular morbidity in women. Adjusting for post-exposure factors that are colliders could result in inflated associations.^54^ VB is known to be associated with obstetric complications such as abruptio placentae and placenta praevia.^55^ We adjusted analyses for preeclampsia-eclampsia, abruptio placentae, and placenta praevia at the index pregnancy, assuming these were not mediators but proxies of the underlying vascular condition potentially also leading to VB, which manifested post-exposure. Results of analyses additionally adjusted vs unadjusted for placental complications did not differ appreciably.

### Strengths and limitations of the study

The strengths of this study are population-based design, up to 25 years of available follow-up in the main analysis, use of validated algorithms for the outcomes identification and nearly complete follow-up of women until emigration, death, or the outcome of interest, which minimizes the selection bias.

The VB diagnosis was not validated in the DNPR. However, one Danish study showed that most women with VB in pregnancy received ultrasound^6^ required to establish foetal heart activity at bleeding presentation and exclude miscarriage and ectopic pregnancy. The number of false-positive VBs in this study is likely small. At the same time, we were unable to capture episodes of VB not resulting in a hospital-based contact and, therefore, could miss a considerable proportion of pregnancies affected by milder bleeding.^6^ Non-differential misclassification of pregnancies’ status would result in diluted associations. The diagnoses for miscarriage (spontaneous abortion) in the DNPR were previously validated and showed PPV of 97%.^56^ The registration of diagnostic and procedure codes for medical and surgical abortions (pregnancy terminations) was not validated in the DNPR, however, the induced abortion diagnosis in the Register of Legally Induced Abortions had PPV of 94% and procedure codes, in general, have high validity in the DNPR.^23^

The use of outcome diagnoses was validated with high PPV for diabetes (82-96%),^57^ hypertension (92%), PCI (98%),^58^ and slightly lower PPV for heart failure (76%),^58^ and haemorrhagic stroke (71-80%).^59,60^ The PPV for ischaemic stroke was 71-85%;^59,60^ however, since the definition of ischaemic stroke in this study included undefined stroke diagnoses, a larger proportion of false positives may be present. Non-differential misclassification of the outcome is expected to dilute the associations toward or beyond the null value.

The residual confounding by obesity is likely since data only on hospital-based diagnoses were available. Although additional adjustment of analyses for BMI did not meaningfully shift the aHR estimates, the concern of unmeasured and residual confounding remains. The prevalence of PCOS was similar across all cohorts in this study, however, we had no data on other indicators of low-grade inflammation in women.

The results of this paper present total associations, including pathways mediated via post-exposure conditions. For instance, the association with heart failure included the path mediated by previous hypertension, myocardial infarction and other ischaemic heart disease. Although we did not investigate the specific pathways of cardiovascular morbidity in women following a VB-affected pregnancy, associations with diabetes type 2 and hypertension suggest that metabolic syndrome may be one of them. We cannot exclude that part of the observed association between VB and the outcomes may be mediated via underlying mechanisms associated with miscarriages of women’s subsequent pregnancies. Association between VB-affected pregnancy and diabetes mellitus type 1 is unlikely to be explained by the same pathways as associations with diabetes mellitus type 2 and may point to the residual confounding. Common metabolic and pro-inflammatory pathways for VB, miscarriage and vascular pathology are possible and may at least partly explain away observed associations.

## CONCLUSION

Having a pregnancy affected by VB before 20 gestational weeks was associated with increased risks of diabetes type 1 and type 2, hypertension, ischaemic heart disease, including myocardial infarction, atrial fibrillation or flutter, heart failure, and ischaemic and haemorrhagic stroke in women when compared with having a VB-unaffected pregnancy or termination, but not miscarriage.

## Data Availability

Data is not available for sharing to protect the identity of participants according to Danish legislation.

## Contributors

ED, EHP, HTS, VE designed the study. HTS facilitated the data application process. ED designed the statistical analysis plan. ED conducted the analyses, EHP provided expertise and supervision of the data analysis. All authors participated in the discussion of the results and their critical interpretation. ED drafted the manuscript, organised the writing, and produced a graphical presentation of the results. HTS provided clinical expertise. All authors equally participated in the critical revision of the manuscript and approved its final version. HTS is the guarantor.

## Funding

Department of Clinical Epidemiology, Aarhus University partaking in studies with institutional funding from regulators and pharmaceutical companies, given as research grants to and administered by Aarhus University. None of these projects is related to the current study.

## Competing interests

All authors report no competing interest

## Ethics approval

not required for registry-based research in Denmark.

## Data sharing

data are not available for sharing to protect the identity of participants according to Danish legislation.

## Notes

### Competing Interest Statement

The authors have declared no competing interest.

### Author Declarations

No patient informed consent or ethical approval is required for registry-based research according to Danish legislation. The study was reported to the Danish Data Protection Agency (according to the Danish legislation) through registration at Aarhus University (record number: AU-2016-051-000001, sequential number 605).

### Summary of Updates

Additional sensitivity analyses for acute MI and stroke as the outcomes were performed.

