## Supplementary materials for "Risk of diabetes and cardiovascular diseases in women with vaginal bleeding before 20 gestational weeks: Danish population-based cohort study"

**Supplemental materials**

[**Table 1. Definitions of the exposure, outcomes, and covariables (ICD-8, ICD-10, and Anatomical Therapeutic Classification [ATC] codes)**	2](#_Toc119427826)

### **Table 1. Definitions of the exposure, outcomes, and covariables (ICD-8, ICD-10, and Anatomical Therapeutic Classification [ATC] codes)**

|  | ICD-8 codes | ICD-10 codes | Procedure codes | ATC codes |
| --- | --- | --- | --- | --- |
| **Exposure and comparators** | | | | |
| Vaginal bleeding in pregnancy (threatened abortion) | 63239 | DO200 | N/A | N/A |
| Miscarriage (spontaneous abortion) | 643; 634;  645 | O03; O021A | N/A | N/A |
| Pregnancy termination (medical or surgical abortion) | 640; 641; 642 | O04 | 94520 (abortus provocatus medicamentalis); KLCH (termination of pregnancy) | N/A |
| **Exclusion criteria diagnoses** | | | | |
| Hyperlipidaemia | 27201, 27208, 27209 | E78.1-E78.5 | N/A | N/A |
| Hypercholesterolemia | 27200 | E78.0 | N/A | N/A |
| Arterial claudication | 44389-44399 | I739A | N/A | N/A |
| **Covariables** | | | | |
| Obesity | 27799 | E66 | N/A | N/A |
| PCOS | 25690 | E28.2 | N/A | N/A |
| Thyroid disorders | 240-246 | E00-E07 | N/A | N/A |
| Rheumatic diseases with heart involvement | 39099, 39199, 39299, 39209, 39300, 39301 | I00.0-I02.9; I05.0-I09.9 | N/A | N/A |
| COPD | 490-493; 515-518 | J40-J47; J60-J67;  J68.4; J70.1; J70.3;  J84.1; J92.0; J96.1;  J98.2; J98.3 | N/A | N/A |
| Chronic kidney diseases | 24902, 25002, 75310-75319, 582, 583, 584, 59009, 59320, 792 | 249.02, 250.02, 753.10-753.19, 582, 583, 584, 590.09, 593.20, 792;  E10.2, E11.2, E14.2,  N03, N05, N11.0,  N14; N16, N18-N19,  N26.9, Q61.1-Q61.4 | N/A | N/A |
| Liver diseases | 571, 57300, 57301, 57304  45600-45609 | K70.0, K70.3, K71.7,  K73, K74, K76.0,  B18, I85 | N/A | N/A |
| Cancer | 140-209 | C00-C96 | N/A | N/A |
| Preeclampsia superimposed on chronic hypertension | N/A | O11 | N/A | N/A |
| Gestational proteinuria | 63701, 63702 | O12 | N/A | N/A |
| Gestational hypertension | 63700 | O13 | N/A | N/A |
| Preeclampsia | 63703, 63704, 63709 | O14 | N/A | N/A |
| Eclampsia | 63719 | O15 | N/A | N/A |
| Any hypertensive disorder of pregnancy | 63701, 63702, 63700, 63703, 63704, 63709, 63719 | O11- O15 | N/A | N/A |
| Placenta praevia | 6510x; 6511x; 6512x; 6513x | O44 | N/A | N/A |
| Abruptio placentae | 6321x; 6514x | O45 |  |  |
| Gestational diabetes | 63474, 6449x, Y6449 | O244, O249 | N/A | N/A |
| Any psychiatric disorder | 290-315 | F00-F99 | N/A | N/A |
| Mood disorders | 296.x9 (excluding 296.89), 298.09, 298.19, 300.49, 301.19 | F30-F39, F92.0 | N/A | N/A |
| Schizophrenia | 295x9, 9689, 297x9, 9829-29899, 29904, 29905, 9909, 30183 | F20-29 | N/A | N/A |
| Autism spectrum disorder | 29901-29903 | F84.0-84.1, F84.5, F84.8-84.9 | N/A | N/A |
| Neurotic disorders | 300x9 (Excluding 30049), 305x9, 30568, 30799 | F40-F48 | N/A | N/A |
| Eating disorders | 30560, 30650, 30658, 30659 | F50 | N/A | N/A |
| Personality disorders | 301x9 (excluding 30119), 30180, 30181, 30182, 30184 | F60 | N/A | N/A |
| Intellectual disabilities | 311, 312, 313, 314, 315 | F70-F79 | N/A | N/A |
| Behavioral disorders | 306x9, 3080x | F90-F98 | N/A | N/A |
| Alcohol or drug abuse | 291x, 303x9, 30320, 30328, 30390, 29439, 304x9; 57710, 57109, 57110 | F10, F11-F19; Z72.1, T51.0, K86.0, G31.2, G62.1, G72.1, I42.6, K29.2, R78.0, Z71.4 | N/A | N/A |
| **Comedications** | | | | |
| Antipsychotics | N/A | N/A | N/A | N05A |
| Mood disorders medication | N/A | N/A | N/A | N06 |
| NSAIDs | N/A | N/A | N/A | M01A, N02BA |
| Thyroid disorders medication | N/A | N/A | N/A | H03 |
| Steroids for systemic use | N/A | N/A | N/A | H02A, H02B |
| Anti-infectives for systemic use | N/A | N/A | N/A | J01, J02, J04, J09, J12 |
| Antiepileptics | N/A | N/A | N/A | N03A |
| ADHD medication | N/A | N/A | N/A | N06BA01, N06BA02, N06BA04, N06BA09, N06BA12 |
| Aspirin | N/A | N/A | N/A | B01AC06 |
| Clopidogrel | N/A | N/A | N/A | B01AC04 |
| Statins | N/A | N/A | N/A | C10AA, C10B |
| **Outcomes** | | | | |
| Diabetes mellitus type 2 with end organ damage conditions | 250 | E11, E891, G590, G632, G730A, G990C, H280, H360, I792A, M142, N083 | N/A | A10B  *The first redeemed dispensing defined the date of diabetes mellitus type 2 outcome if no prior diagnostic code for diabetes mellitus type 2 was recorded. Otherwise, the date associated with the diagnostic record for diabetes mellitus type 2 defined the date of the outcome. |
| Hypertension | 40009-40499 | I10.0-I15.9 | N/A | C02A, C02B, C02C (alpha-blockers); C02DA, C02L, C03A, C03B, C03D, C03E, C03X, C07C, C07D, C08G, C09BA, C09DA, C09XA52 (non-loop diuretics); C02BD, C02DD, C02DG, C04, C05 (vasodilators); C07 (beta-blockers); C08, C07F, C09BB, C09DB (calcium channel blockers); C09 (renin-angiotensin system [RAS]-acting agents: ACE inhibitors and ARBs).*  *The second of 2+ redeemed dispensings of different antihypertensive drug classes within 180 days of each other defined the date of hypertension outcome if no prior diagnostic code for hypertension was recorded. Otherwise, the date associated with the diagnostic record for hypertension defined the date of the outcome. |
| Ischaemic heart disease | 410-414 | I20-I25 | N/A | N/A |
| Myocardial infarction | 410 | I21 | N/A | N/A |
| Heart failure | 42709-42719 | I110, I130, I132, I50 | N/A | N/A |
| Ischaemic stroke | 433-434 | I63-I64 | N/A | N/A |
| Haemorrhagic stroke | 430-432 | I60, I61 | N/A | N/A |
| Atrial fibrillation | 42793, 42794 | I48 | N/A | N/A |
| **Other exclusion criteria before the index date** | | | | |
| Diabetes mellitus type 1 (same codes are used for diabetes mellitus type 1 and 2 until 1987^1^) | 249 | E10, O24.0 | N/A | A10A*  *The first redeemed dispensing defined the date of diabetes mellitus type 1 if no prior diagnostic code for diabetes mellitus type 1 was recorded. Otherwise, the date associated with the diagnostic record for diabetes mellitus type 1 was used. |
| **Additional outcomes** |  |  |  |  |
| Coronary artery bypass graft (CABG) | N/A | N/A | 30009, 30019, 30029, 30039, 30049, 30059, 30069, 30079, 30089, 30099, 30109, 30119, 30120, 30129, 30139, 30149, 30159, 30169, 30179, 30189, 30199, 30200, KFNA-KFNE, KFNH20 | N/A |
| Percutaneous coronary intervention (PCI) | N/A | N/A | 30350, 30354, 30240, KFNG, KFNF | N/A |

### **Figure 1. Directed acyclic graph (DAG) depicting the relationship between measured and unmeasured variables in the study**

**Abbreviations**: PREG, pregnancy; PREG_BMI_t0_, body-mass index at the index pregnancy (at time zero); PRE**-**t0**,** marks covariables measured before the index date (before time zero); TAB, threatened abortion; U_1_, an unmeasured shared cause for both miscarriage and pregnancy termination and the OUTCOME_t0+fup_; U_2_, an unmeasured factor increasing the risk of death and of the OUTCOME_t0+fup_ (death is a competing event and the outcomes are only observed among those living “DEATH=0”); FERTILITY is unmeasured (latent node)

**Exposure**: TAB-affected pregnancy within the first 20 gestational weeks (PREG_childbirthTAB_t0_)

**Outcome**: cardiovascular events (diabetes mellitus type 1 and 2, hypertension, myocardial infarction, ischaemic heart disease, heart failure, atrial fibrillation or flutter, ischaemic stroke, haemorrhagic stroke, CABG, PCI)

Directed solid line arrows: causal relationship (including temporality, e.g. A → B means that A causes and precedes B)

Bi-directed dashed line arrows: associations confounded by the unmeasured common shared causes (A ⟷ B means A ←U→ B, where U is unobserved)

DAG tool: https://causalfusion.net/app^2^

### **Computation of inverse probability of treatment weights (IPTW) for average treatment effect (ATE) estimation**

We computed stabilized IPTW as the ratio of the probability of being exposed, i.e. having vaginal bleeding within 20 gestational weeks (Pr[A=1]), and the probability of not being exposed (1-Pr[A=1]) for comparison cohorts in the numerator and conditional probability of exposure level observed given the covariates in the denominator, i.e. Pr[A=1|L] for VB-affected pregnancies and 1-Pr[A=1|L] otherwise.^3–7^

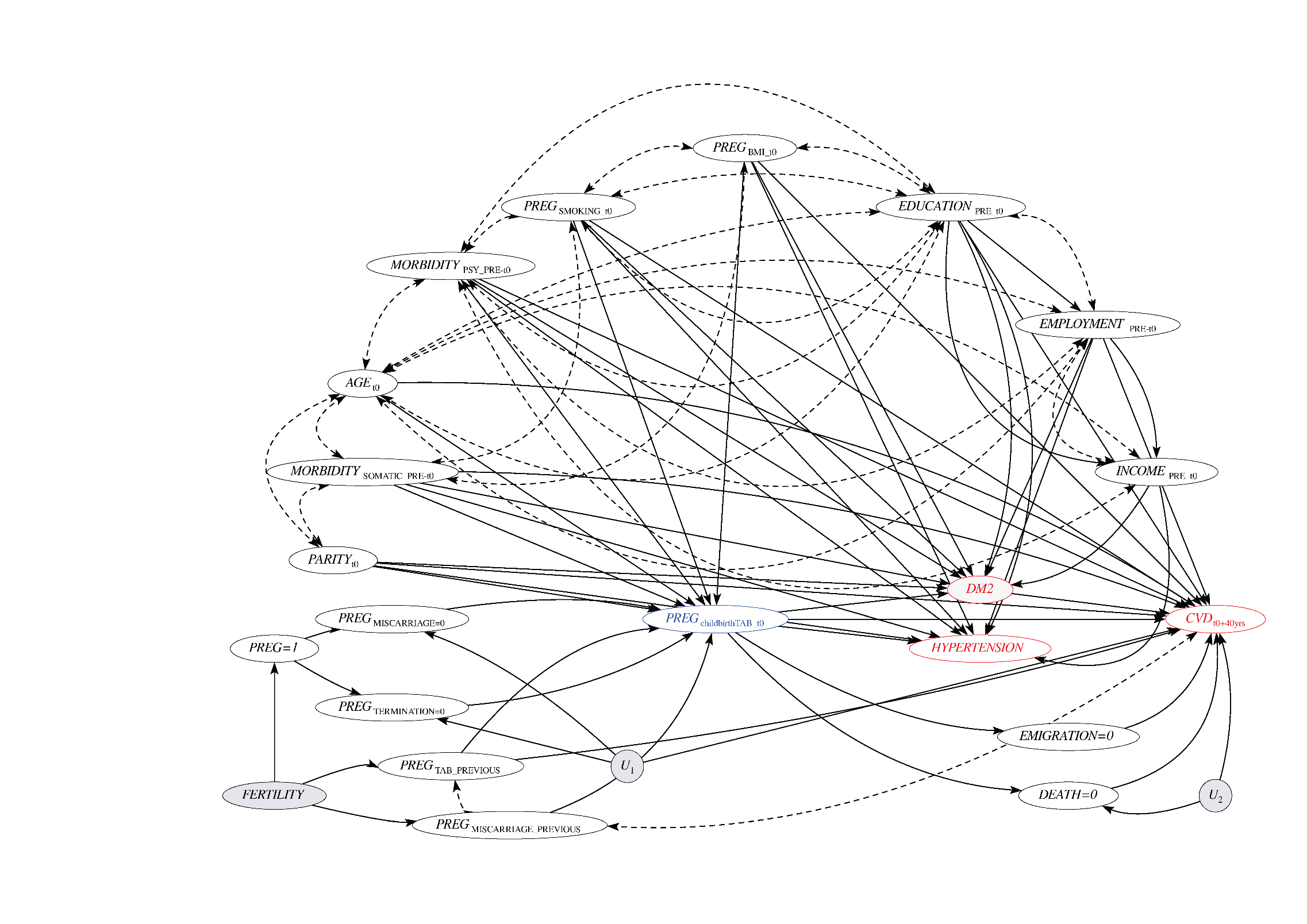

### **Figure 2. Cumulative incidence curves for diabetes and cardiovascular diseases in women following vaginal bleeding (VB)-affected and VB-unaffected pregnancy, termination or miscarriage, Denmark, 1994-2018**

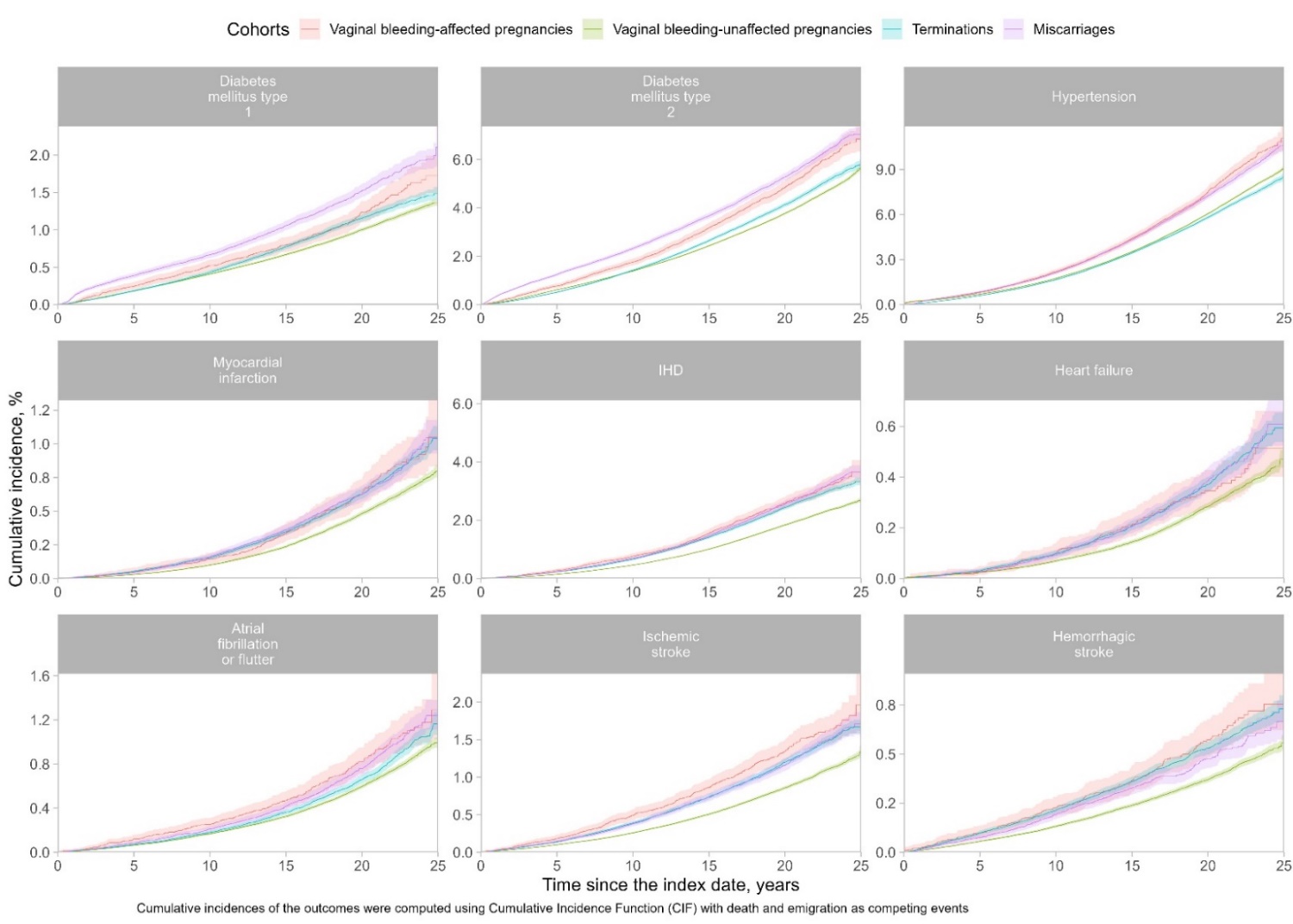

IHD, ischemic heart disease

### **Table 2. Cumulative incidences of diabetes and cardiovascular diseases in women following vaginal bleeding (VB)-affected and VB-unaffected pregnancy, termination or miscarriage, Denmark, 1994-2018**

| **Outcome** | **Time, years** | **Cumulative incidence, % (95% CI)** | | | |
| --- | --- | --- | --- | --- | --- |
|  |  | **VB-affected pregnancy** | **VB-unaffected pregnancy** | **Termination** | **Miscarriage** |
| Diabetes mellitus type 1 | 5 | 0.2 (0.2-0.3) | 0.2 (0.2-0.2) | 0.2 (0.2-0.2) | 0.4 (0.4-0.4) |
|  | 10 | 0.5 (0.5-0.6) | 0.4 (0.4-0.4) | 0.4 (0.4-0.5) | 0.7 (0.6-0.7) |
|  | 20 | 1.2 (1.1-1.4) | 1.0 (1.0-1.0) | 1.2 (1.1-1.2) | 1.5 (1.4-1.6) |
|  | 25 | 1.7 (1.5-2.0) | 1.4 (1.3-1.4) | 1.5 (1.4-1.6) | 2.1 (1.9-2.4) |
| Diabetes mellitus type 2 | 5 | 0.8 (0.7-0.9) | 0.6 (0.6-0.6) | 0.5 (0.5-0.5) | 1.2 (1.2-1.3) |
|  | 10 | 1.8 (1.6-1.9) | 1.4 (1.4-1.4) | 1.4 (1.4-1.5) | 2.3 (2.2-2.4) |
|  | 20 | 4.8 (4.6-5.1) | 3.8 (3.7-3.8) | 4.1 (4.0-4.2) | 5.3 (5.1-5.4) |
|  | 25 | 6.9 (6.3-7.4) | 5.6 (5.5-5.8) | 5.8 (5.6-6.0) | 7.0 (6.8-7.3) |
| Hypertension | 5 | 0.8 (0.7-0.9) | 0.7 (0.7-0.7) | 0.6 (0.6-0.6) | 0.8 (0.8-0.9) |
|  | 10 | 2.2 (2.0-2.4) | 1.8 (1.7-1.8) | 1.7 (1.6-1.7) | 2.1 (2.1-2.2) |
|  | 20 | 7.5 (7.1-7.8) | 6.1 (6.0-6.1) | 5.8 (5.7-5.9) | 7.3 (7.1-7.5) |
|  | 25 | 11.1 (10.3-11.9) | 9.0 (8.9-9.2) | 8.5 (8.2-8.8) | 10.6 (10.2-11.0) |
| Ischemic heart disease | 5 | 0.3 (0.2-0.3) | 0.1 (0.1-0.2) | 0.2 (0.2-0.2) | 0.2 (0.2-0.3) |
|  | 10 | 0.8 (0.7-0.9) | 0.5 (0.4-0.5) | 0.7 (0.6-0.7) | 0.7 (0.6-0.7) |
|  | 20 | 2.6 (2.4-2.8) | 1.8 (1.8-1.9) | 2.4 (2.4-2.5) | 2.6 (2.4-2.7) |
|  | 25 | 3.6 (3.3-4.1) | 2.7 (2.6-2.7) | 3.3 (3.2-3.5) | 4.4 (3.2-6.2) |
| Myocardial infarction | 5 | 0.1 (0.0-0.1) | 0.0 (0.0-0.0) | 0.1 (0.0-0.1) | 0.0 (0.0-0.1) |
|  | 10 | 0.1 (0.1-0.2) | 0.1 (0.1-0.1) | 0.2 (0.1-0.2) | 0.2 (0.1-0.2) |
|  | 20 | 0.6 (0.5-0.7) | 0.5 (0.5-0.5) | 0.6 (0.6-0.7) | 0.6 (0.6-0.7) |
|  | 25 | 1.1 (0.8-1.3) | 0.8 (0.8-0.8) | 1.0 (1.0-1.1) | 1.0 (0.9-1.2) |
| Atrial fibrillation or flutter | 5 | 0.1 (0.1-0.2) | 0.1 (0.1-0.1) | 0.1 (0.1-0.1) | 0.1 (0.1-0.1) |
|  | 10 | 0.3 (0.2-0.3) | 0.2 (0.2-0.2) | 0.2 (0.2-0.2) | 0.2 (0.2-0.2) |
|  | 20 | 0.8 (0.7-0.9) | 0.6 (0.6-0.6) | 0.7 (0.6-0.7) | 0.8 (0.7-0.8) |
|  | 25 | 1.3 (1.0-1.6) | 1.0 (0.9-1.0) | 1.2 (1.1-1.3) | 1.2 (1.1-1.4) |
| Heart failure | 5 | 0.0 (0.0-0.0) | 0.0 (0.0-0.0) | 0.0 (0.0-0.0) | 0.0 (0.0-0.0) |
|  | 10 | 0.1 (0.1-0.1) | 0.1 (0.1-0.1) | 0.1 (0.1-0.1) | 0.1 (0.1-0.1) |
|  | 20 | 0.3 (0.3-0.4) | 0.3 (0.3-0.3) | 0.4 (0.3-0.4) | 0.4 (0.3-0.4) |
|  | 25 | 0.5 (0.4-0.7) | 0.5 (0.4-0.5) | 0.6 (0.5-0.7) | 0.6 (0.5-0.7) |
| Ischemic stroke | 5 | 0.2 (0.1-0.2) | 0.1 (0.1-0.1) | 0.1 (0.1-0.2) | 0.1 (0.1-0.2) |
|  | 10 | 0.5 (0.4-0.6) | 0.3 (0.2-0.3) | 0.4 (0.4-0.4) | 0.4 (0.3-0.4) |
|  | 20 | 1.3 (1.2-1.5) | 0.9 (0.8-0.9) | 1.2 (1.1-1.3) | 1.2 (1.1-1.2) |
|  | 25 | 2.0 (1.6-2.4) | 1.3 (1.3-1.4) | 1.7 (1.6-1.8) | 1.7 (1.6-1.9) |
| Haemorrhagic stroke | 5 | 0.1 (0.1-0.1) | 0.1 (0.1-0.1) | 0.1 (0.1-0.1) | 0.1 (0.1-0.1) |
|  | 10 | 0.2 (0.2-0.3) | 0.1 (0.1-0.1) | 0.2 (0.2-0.2) | 0.2 (0.2-0.2) |
|  | 20 | 0.6 (0.5-0.7) | 0.4 (0.4-0.4) | 0.5 (0.5-0.6) | 0.5 (0.4-0.5) |
|  | 25 | 0.8 (0.6-0.9) | 0.6 (0.5-0.6) | 0.7 (0.7-0.8) | 0.7 (0.6-0.8) |

### **Table 3. Conventionally adjusted hazard ratios (HR) with 95% confidence intervals (CI) for diabetes and cardiovascular diseases in women with VB-affected vs comparators, Denmark, 1994-2018**

|  |  | **Pregnancy observations (1994-2017)** | |
| --- | --- | --- | --- |
| **Outcomes** | **VB-affected pregnancy vs Comparator** | **All observations^a^** | **First observations^b^** |
|  |  | **HR (95% CI)^c^** | |
| **Diabetes mellitus type 1** | VB-unaffected pregnancy^d^ | 1.08 (0.97-1.20) | 1.16 (0.95-1.42) |
|  | Termination | 1.22 (1.08-1.38) | 1.17 (0.94-1.45) |
|  | Miscarriage | 0.83 (0.73-0.95) | 0.72 (0.57-0.89) |
| **Diabetes mellitus type 2** | VB-unaffected pregnancy^d^ | 1.15 (1.09-1.22) | 1.29 (1.16-1.42) |
|  | Termination | 1.29 (1.21-1.37) | 1.26 (1.13-1.40) |
|  | Miscarriage | 0.86 (0.81-0.93) | 0.80 (0.72-0.90) |
| **Hypertension** | VB-unaffected pregnancy^d^ | 1.15 (1.10-1.20) | 1.15 (1.07-1.24) |
|  | Termination | 1.25 (1.19-1.31) | 1.68 (1.55-1.83) |
|  | Miscarriage | 1.11 (1.05-1.18) | 1.07 (0.98-1.17) |
| **Ischemic heart disease** | VB-unaffected pregnancy^d^ | 1.19 (1.11-1.29) | 1.14 (0.95-1.36) |
|  | Termination | 1.11 (1.02-1.20) | 1.36 (1.12-1.65) |
|  | Miscarriage | 1.08 (0.97-1.19) | 1.03 (0.84-1.27) |
| **Myocardial infarction** | VB-unaffected pregnancy^d^ | 1.11 (0.95-1.29) | 1.32 (0.93-1.86) |
|  | Termination | 1.01 (0.85-1.20) | 1.53 (1.06-2.21) |
|  | Miscarriage | 1.00 (0.81-1.23) | 1.33 (0.89-1.99) |
| **Atrial fibrillation or flutter** | VB-unaffected pregnancy^d^ | 1.23 (1.08-1.40) | 1.45 (1.12-1.88) |
|  | Termination | 1.27 (1.09-1.47) | 1.90 (1.43-2.53) |
|  | Miscarriage | 1.13 (0.94-1.35) | 1.23 (0.91-1.66) |
| **Heart failure** | VB-unaffected pregnancy^d^ | 1.08 (0.88-1.32) | 0.98 (0.62-1.56) |
|  | Termination | 1.00 (0.80-1.25) | 1.19 (0.73-1.93) |
|  | Miscarriage | 1.05 (0.80-1.37) | 1.15 (0.68-1.96) |
| **Ischemic stroke** | VB-unaffected pregnancy^d^ | 1.36 (1.23-1.51) | 1.17 (0.92-1.50) |
|  | Termination | 1.16 (1.03-1.30) | 1.22 (0.94-1.58) |
|  | Miscarriage | 1.03 (0.89-1.19) | 0.97 (0.74-1.29) |
| **Hemorrhagic stroke** | VB-unaffected pregnancy^d^ | 1.31 (1.12-1.53) | 1.41 (1.03-1.95) |
|  | Termination | 1.09 (0.91-1.30) | 1.46 (1.04-2.04) |
|  | Miscarriage | 1.18 (0.95-1.46) | 1.35 (0.93-1.96) |

^a^ Different pregnancies of the same woman were included and could contribute person-time to different cohorts.

^b^ We followed the first pregnancy of each woman. The cohort membership was classified according to the exposure status at the first pregnancy identifiable via the registries.

^c^ Adjusted using IPT-weighted Cox proportional hazards regression for women’s age at pregnancy end, calendar year, parity, civil status, employment, highest attained education, income in year-specific quartiles, reproductive history (history of at least one previous vaginal bleeding, termination, miscarriage), comorbidities (chronic pulmonary disease, chronic kidney disease, chronic liver disease, cancer, rheumatic conditions, thyroid conditions, alcohol abuse, and individual classes of psychiatric conditions) and medication use (antipsychotics, medication for mood disorders, NSAIDs, aspirin, thyroid disorders medication, steroids for systemic use, anti-infectives for systemic use).

^d^ Analyses were additionally adjusted for smoking status recorded at 1st prenatal visit and for preeclampsia-eclampsia, placenta praevia, and abruptio placentae at the delivery of the index pregnancy.

### **Table 4. Sensitivity analysis 1: Adjusted hazard ratios (HR) with 95% confidence intervals (CI) for diabetes and cardiovascular diseases in women with VB-affected vs comparators, Denmark, 1979-2018**

|  |  | **Pregnancy observations (1979-1993)** | |
| --- | --- | --- | --- |
| **Outcomes** | **VB-affected pregnancy vs Comparator** | **All observations^a^** | **First observations^b^** |
|  |  | **HR (95% CI)^c^** | |
| **Diabetes mellitus type 1** | VB-unaffected pregnancy^d^ | 1.16 (1.07-1.25) | 1.16 (1.02-1.32) |
|  | Termination | 1.23 (1.11-1.36) | 1.29 (1.10-1.52) |
|  | Miscarriage | 0.92 (0.83-1.02) | 0.97 (0.84-1.13) |
| **Diabetes mellitus type 2** | VB-unaffected pregnancy^d^ | 1.21 (1.16-1.27) | 1.31 (1.22-1.41) |
|  | Termination | 1.23 (1.16-1.31) | 1.35 (1.23-1.48) |
|  | Miscarriage | 1.03 (0.97-1.09) | 1.09 (1.01-1.19) |
| **Hypertension** | VB-unaffected pregnancy^d^ | 1.16 (1.13-1.20) | 1.14 (1.09-1.19) |
|  | Termination | 1.11 (1.07-1.15) | 1.16 (1.09-1.22) |
|  | Miscarriage | 1.04 (0.99-1.08) | 1.02 (0.97-1.07) |
| **Ischemic heart disease** | VB-unaffected pregnancy^d^ | 1.46 (1.39-1.53) | 1.45 (1.34-1.57) |
|  | Termination | 1.23 (1.16-1.31) | 1.37 (1.24-1.50) |
|  | Miscarriage | 1.13 (1.06-1.20) | 1.21 (1.10-1.32) |
| **Myocardial infarction** | VB-unaffected pregnancy^d^ | 1.33 (1.21-1.46) | 1.25 (1.06-1.47) |
|  | Termination | 1.05 (0.93-1.18) | 1.12 (0.92-1.36) |
|  | Miscarriage | 0.95 (0.83-1.08) | 1.02 (0.86-1.22) |
| **Atrial fibrillation or flutter** | VB-unaffected pregnancy^d^ | 1.25 (1.16-1.36) | 1.24 (1.08-1.41) |
|  | Termination | 1.03 (0.92-1.14) | 0.99 (0.85-1.17) |
|  | Miscarriage | 0.95 (0.85-1.06) | 0.95 (0.82-1.10) |
| **Heart failure** | VB-unaffected pregnancy^d^ | 1.34 (1.20-1.50) | 1.29 (1.07-1.56) |
|  | Termination | 0.96 (0.83-1.11) | 1.03 (0.83-1.29) |
|  | Miscarriage | 0.92 (0.79-1.08) | 0.97 (0.79-1.19) |
| **Ischemic stroke** | VB-unaffected pregnancy^d^ | 1.33 (1.23-1.43) | 1.35 (1.20-1.53) |
|  | Termination | 1.08 (0.99-1.19) | 1.13 (0.97-1.31) |
|  | Miscarriage | 1.01 (0.92-1.12) | 1.09 (0.95-1.25) |
| **Hemorrhagic stroke** | VB-unaffected pregnancy^d^ | 1.32 (1.18-1.48) | 1.23 (1.01-1.49) |
|  | Termination | 1.09 (0.94-1.26) | 1.06 (0.84-1.36) |
|  | Miscarriage | 1.04 (0.89-1.22) | 1.05 (0.84-1.30) |
| **Additional CVD outcomes** | | | |
| **CABG** | VB-unaffected pregnancy^d^ | 1.28 (0.98-1.68) | 1.44 (0.94-2.20) |
|  | Termination | 0.95 (0.67-1.35) | 1.12 (0.69-1.80) |
|  | Miscarriage | 0.93 (0.63-1.37) | 1.02 (0.64-1.62) |
| **PCI** | VB-unaffected pregnancy^d^ | 1.20 (1.06-1.35) | 1.23 (1.01-1.49) |
|  | Termination | 1.01 (0.87-1.18) | 1.17 (0.92-1.47) |
|  | Miscarriage | 0.94 (0.80-1.11) | 1.05 (0.85-1.31) |

CABG, coronary artery bypass graft; PCI, percutaneous coronary intervention

^a^ Different pregnancies of the same woman were included and could contribute person-time to different cohorts.

^b^ We followed a first pregnancy of each woman identifiable via the MBR and the DNPR. The cohort membership was classified according to the exposure status at the first pregnancy.

^c^ Adjusted using IPT-weighted Cox proportional hazards regression for women’s age at pregnancy end, calendar year, parity, number of previous pregnancies ending in childbirth, termination or miscarriage identifiable via the MBR and the DNPR, civil status, employment, highest attained education, income in year-specific quartiles, reproductive history (histories of at least one previous vaginal bleeding, termination, miscarriage, preeclampsia-eclampsia, placenta praevia, abruption placentae), comorbidities (obesity, chronic pulmonary disease, chronic kidney disease, chronic liver disease, cancer, rheumatic conditions, thyroid conditions, alcohol abuse).

^d^ Analyses were additionally adjusted for preeclampsia-eclampsia, placenta praevia, and abruption placentae at the delivery of the index pregnancy.

### **Table 5. Sensitivity analysis 2: Adjusted hazard ratios (HR) with 95% confidence intervals (CI) for diabetes and cardiovascular diseases in women with VB-affected vs comparators Denmark, 2004-2018**

|  |  | **Pregnancy observations (2004-2017)** | |
| --- | --- | --- | --- |
| **Outcomes** | **VB-affected pregnancy vs Comparator** | **All observations^a^** | **First observations^b^** |
|  |  | **HR (95% CI)^c^** | |
| **Diabetes mellitus type 1** | VB-unaffected pregnancy^d^ | 1.19 (0.94-1.51) | 1.02 (0.64-1.64) |
|  | Termination | 1.37 (1.00-1.89) | 1.51 (0.82-2.80) |
|  | Miscarriage | 0.76 (0.56-1.04) | 0.50 (0.29-0.83) |
| **Diabetes mellitus type 2** | VB-unaffected pregnancy^d^ | 1.29 (1.15-1.45) | 1.38 (1.13-1.68) |
|  | Termination | 1.28 (1.08-1.51) | 1.33 (1.00-1.78) |
|  | Miscarriage | 0.74 (0.64-0.87) | 0.61 (0.48-0.77) |
| **Hypertension** | VB-unaffected pregnancy^d^ | 1.33 (1.19-1.48) | 1.53 (1.30-1.80) |
|  | Termination | 1.47 (1.26-1.71) | 2.16 (1.70-2.74) |
|  | Miscarriage | 1.25 (1.08-1.44) | 1.16 (0.95-1.40) |
| **Ischemic heart disease** | VB-unaffected pregnancy^d^ | 1.54 (1.25-1.88) | 1.24 (0.75-2.06) |
|  | Termination | 1.25 (0.93-1.67) | 0.92 (0.46-1.88) |
|  | Miscarriage | 1.20 (0.90-1.59) | 1.09 (0.60-1.98) |
| **Myocardial infarction** | VB-unaffected pregnancy^d^ | 1.38 (0.89-2.14) | 1.56 (0.56-4.34) |
|  | Termination | 0.87 (0.44-1.73) | 0.72 (0.21-2.49) |
|  | Miscarriage | 0.96 (0.53-1.75) | 1.04 (0.33-3.27) |
| **Atrial fibrillation or flutter** | VB-unaffected pregnancy^d^ | 1.50 (1.07-2.09) | 2.14 (1.23-3.73) |
|  | Termination | 1.76 (1.12-2.79) | 3.03 (1.42-6.46) |
|  | Miscarriage | 1.52 (0.97-2.38) | 1.57 (0.82-3.01) |
| **Heart failure** | VB-unaffected pregnancy^d^ | 1.25 (0.72-2.18) | 1.08 (0.34-3.42) |
|  | Termination | 1.21 (0.62-2.35) | 1.97 (0.56-6.86) |
|  | Miscarriage | 1.08 (0.52-2.25) | 0.94 (0.24-3.73) |
| **Ischemic stroke** | VB-unaffected pregnancy^d^ | 1.39 (1.05-1.83) | 0.93 (0.48-1.84) |
|  | Termination | 0.89 (0.59-1.34) | 0.80 (0.35-1.81) |
|  | Miscarriage | 0.79 (0.53-1.17) | 0.81 (0.38-1.74) |
| **Hemorrhagic stroke** | VB-unaffected pregnancy^d^ | 1.60 (1.07-2.38) | 1.91 (0.92-3.95) |
|  | Termination | 1.29 (0.73-2.28) | 2.02 (0.72-5.67) |
|  | Miscarriage | 0.93 (0.54-1.60) | 1.42 (0.59-3.38) |

Coronary artery bypass graft and percutaneous coronary intervention were not investigated as the outcomes due to insufficient follow-up duration to capture events.

^a^ Different pregnancies of the same woman were included and could contribute person-time to different cohorts.

^b^ We followed a first pregnancy of each woman identifiable via the MBR and the DNPR. The cohort membership was classified according to the exposure status at the first pregnancy.

^c^ Adjusted using IPT-weighted Cox proportional hazards regression for women’s age at pregnancy end, calendar year, parity, number of previous pregnancies ending in childbirth, termination or miscarriage identifiable via registries, civil status, employment, highest attained education, income in year-specific quartiles, reproductive history (histories of at least one previous vaginal bleeding, termination, miscarriage, preeclampsia-eclampsia, placenta praevia, abruption placentae), comorbidities (obesity, chronic pulmonary disease, chronic kidney disease, chronic liver disease, cancer, rheumatic conditions, thyroid conditions, alcohol abuse), history of any psychiatric condition, and medication use (antipsychotics, antiepileptics, medication for mood disorders, NSAIDs or aspirin, thyroid disorders medication, steroids for systemic use, anti-infectives for systemic use).

^d^ Analyses were additionally adjusted for smoking status recorded at 1st prenatal visit, BMI, and preeclampsia-eclampsia, placenta praevia, and abruption placentae at the delivery of the index pregnancy.

### **Table 6. Sensitivity analysis 3: Adjusted hazard ratios (HR) with 95% confidence intervals (CI) for acute myocardial infarction, ischemic and hemorrhagic stroke**

| **Outcomes**^a^ | **VB-affected pregnancy vs Comparator** | **HR (95% CI)^b, c^** |
| --- | --- | --- |
| **Myocardial infarction** | VB-unaffected pregnancy^d^ | 1.47 (1.11-1.96) |
|  | Termination | 1.20 (0.82-1.75) |
|  | Miscarriage | 1.21 (0.81-1.79) |
| **Ischemic stroke** | VB-unaffected pregnancy^d^ | 1.49 (1.25-1.78) |
|  | Termination | 1.04 (0.82-1.33) |
|  | Miscarriage | 0.92 (0.70-1.21) |
| **Hemorrhagic stroke** | VB-unaffected pregnancy^d^ | 1.48 (1.12-1.94) |
|  | Termination | 0.94 (0.64-1.36) |
|  | Miscarriage | 0.93 (0.63-1.36) |
| ^a^ The outcomes were defined using inpatient diagnoses with acute admission type via the Danish National Patient Registry  Different pregnancies of the same woman were included and could contribute person-time to different cohorts.  ^b^ Adjusted using IPT-weighted Cox proportional hazards regression for women’s age at pregnancy end, calendar year, parity, number of previous pregnancies ending in childbirth, termination or miscarriage identifiable via registries, civil status, employment, highest attained education, income in year-specific quartiles, reproductive history (histories of at least one previous vaginal bleeding, termination, miscarriage, preeclampsia-eclampsia, placenta praevia, abruption placentae), comorbidities (obesity, chronic pulmonary disease, chronic kidney disease, chronic liver disease, cancer, rheumatic conditions, thyroid conditions, alcohol abuse), history of any psychiatric condition, and medication use (antipsychotics, antiepileptics, medication for mood disorders, NSAIDs or aspirin, thyroid disorders medication, steroids for systemic use, anti-infectives for systemic use).  ^c^ Analyses were additionally adjusted for smoking status recorded at 1st prenatal visit, BMI, and preeclampsia-eclampsia, placenta praevia, and abruption placentae at the delivery of the index pregnancy. | | |

### **Table 7. E-value plots for the unmeasured confounder or several confounders acting in the same direction associated with vaginal bleeding and with diabetes and cardiovascular diseases beyond measured covariables^8^**

| Comparator: VB-unaffected pregnancy  Outcome: Diabetes mellitus type 1  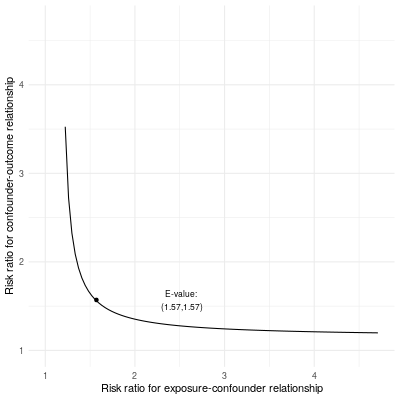 | Comparator: VB-unaffected pregnancy  Outcome: Diabetes mellitus type 2  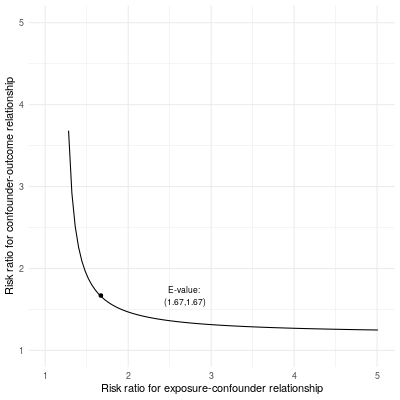 |
| --- | --- |
| Comparator: VB-unaffected pregnancy  Outcome: Hypertension  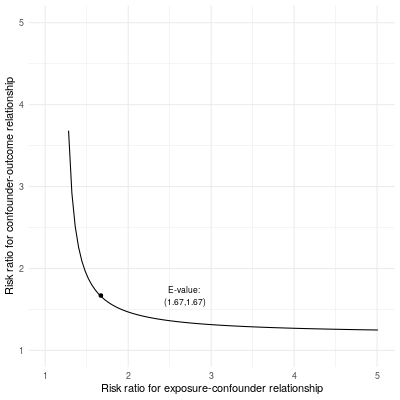 | **Comparator: VB-unaffected pregnancy**  **Outcome: Ischemic heart disease**  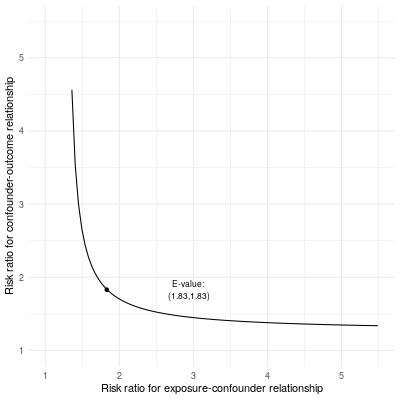 |
| Comparator: VB-unaffected pregnancy  Outcome: Myocardial infarction  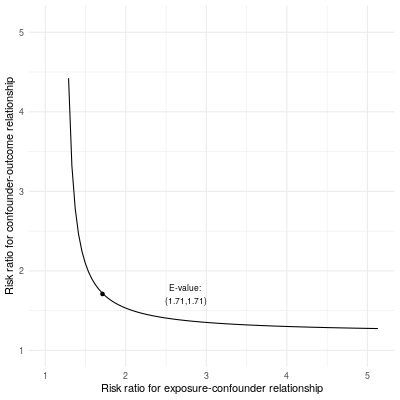 | **Comparator: VB-unaffected pregnancy**  **Outcome: Atrial fibrillation or flutter**  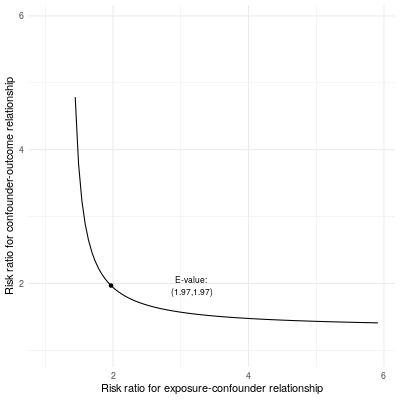 |
| Comparator: VB-unaffected pregnancy  Outcome: Heart failure  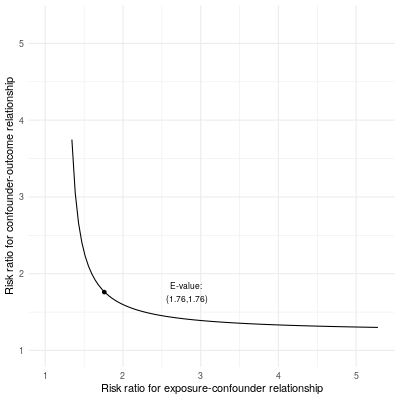 | **Comparator: VB-unaffected pregnancy**  **Outcome: Ischemic stroke**  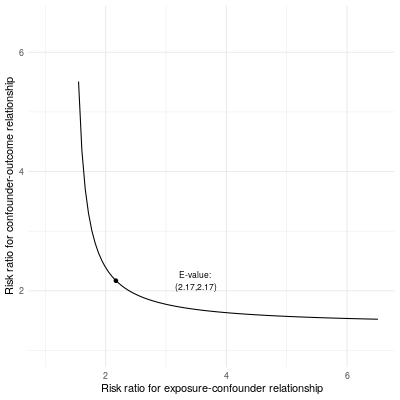 |
| Comparator: VB-unaffected pregnancy  Outcome: Hemorrhagic stroke  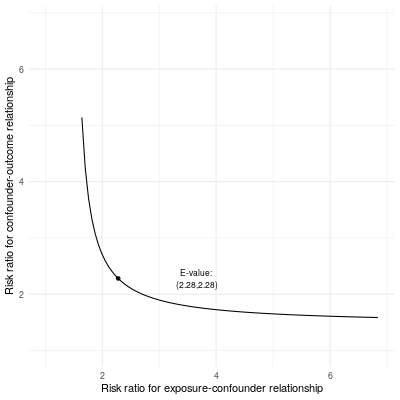 |  |
